# Smooth Curves, Similar Conclusions? Comparing Linear Regression and GAMLSS Neuropsychological Norms

**DOI:** 10.64898/2026.09.01.26361590

**Authors:** Bjørn-Eivind Kirsebom, Ingrid Myrvoll Lorentzen, Jacob Espenes, Ingvild Vøllo Eliassen, Fernando Gonzalez-Ortiz, Anders Wallin, Knut Waterloo, Marie Eckerström, Gøril Rolfseng Grøntvedt, Erik Hessen, Tormod Fladby

## Abstract

**Objective:** Regression-based normative approaches are widely used in neuropsychology but often rely on score transformations to satisfy model assumptions. We compared previously published linear regression (LR)-based norms with norms derived using Generalized Additive Models for Location, Scale and Shape (GAMLSS) for the brief cognitive battery used in the Norwegian Dementia Disease Initiation (DDI) cohort.

**Method:** GAMLSS norms were developed using the same normative samples as the original LR norms for the Consortium to Establish a Registry for Alzheimer’s Disease (CERAD) word list test, Trail Making Test (TMT) A and B, FAS phonemic fluency, and Visual Object and Space Perception Battery (VOSP) Silhouettes. Expected low-score frequencies and empirical base rates were assessed in a normative subsample (n = 131). Clinical implications were evaluated in the DDI clinical cohort (n = 643) using Mild Cognitive Impairment (MCI) classification, two-year diagnostic stability and change, and cerebrospinal fluid (CSF) biomarkers.

**Results:** Compared with LR norms, GAMLSS yielded lower frequencies of low scores, primarily driven by CERAD delayed recall. Nevertheless, concordance between approaches was high (κ = 0.91), with only 4.2% discordant classifications. Two-year diagnostic stability and change were broadly similar across approaches, and CSF biomarker profiles did not clearly favor either normative method.

**Conclusions:** GAMLSS provided a more faithful representation of neuropsychological score distributions, particularly for bounded and non-normal outcomes. However, downstream clinical differences were modest in this setting, suggesting that well-calibrated LR norms may remain robust for clinical classification.

## Introduction

Neuropsychological assessment is central to evaluating severity of neurodegenerative disease, especially for distinguishing mild cognitive impairment (MCI) from normal aging, and interpretation depends on appropriate normative data (Hessen, 2025). However, for several tests commonly used in dementia research and clinical assessment in Scandinavia, available norms have mainly been based on North American samples, and many are now outdated (Ryder, 2021). Over the past decade, our group has sought to address these shortcomings by developing local norms for several neuropsychological measures. This has included a particular focus on normative reference data for younger older adults (>40 years) linked to the Norwegian multicenter Dementia Disease Initiation (DDI) study (Fladby et al., 2017; Kirsebom et al., 2019; Espenes et al., 2020; Eliassen et al., 2020; Lorentzen et al., 2023), which was designed to include and follow individuals in the preclinical and prodromal phases of neurodegenerative disease (Fladby et al., 2017).

In this work, we chose ordinary least squares linear regression (LR), as it offered clear advantages over traditional discrete norms, including more efficient use of sample data and more precise adjustment for demographic influences (Oosterhuis et al., 2016). However, LR relies on distributional assumptions that are not always met by neuropsychological test scores, particularly for bounded or skewed outcomes (Timmerman et al., 2021). In our previous work, most tests therefore required normalization procedures to achieve acceptable residual distributions and satisfy model assumptions. Specifically, raw scores were transformed using cumulative percentile ranks and converted to standardized scaled scores. This approach follows established neuropsychological norming practice and yields clinically interpretable scores (Testa et al., 2009; Crawford et al., 2009), but it also reduces raw-score resolution by grouping adjacent raw scores into coarser scaled-score categories. This was particularly relevant for skewed time-to-completion measures such as the Trail Making Test (TMT) (Espenes et al., 2020) and for bounded or mildly ceilinged score distributions, such as the Consortium to Establish a Registry for Alzheimer’s Disease (CERAD) word list test (WLT) measures (Kirsebom et al., 2019). Although these procedures resulted in acceptable regression models, they also highlighted a broader limitation of conventional regression-based norming: outcome distributions are often reshaped to better fit the model, rather than modeled directly.

Generalized Additive Models for Location, Scale and Shape (GAMLSS) offer a flexible alternative by allowing the distribution of test scores to be modelled more directly, including skewness, kurtosis, and heteroscedasticity (Stasinopoulos et al., 2017; Rigby et al., 2019). This approach has been proposed as particularly relevant for neuropsychological test data, where score distributions often do not fit well with the assumptions underlying LR (Timmerman et al., 2021; Urban et al., 2025). Rather than relying on transformations to make the data approximate normality, GAMLSS makes it possible to choose distributions that better reflect the observed characteristics of the outcome (Rigby et al., 2019). However, despite these methodological advantages, direct comparisons between GAMLSS- and linear regression-based norms remain scarce (Urban et al., 2025). Consequently, it is unclear whether improved statistical model fit translates into meaningful differences in normative calibration, cognitive classification, or other clinically relevant outcomes.

Here, we revisited our previously published regression-based norms for the DDI brief battery using the same normative samples, but within a GAMLSS framework. Our primary aim was to determine whether this more flexible approach provided better normative calibration than the previous LR-based norms. To address this, we compared the two approaches with regard to expected low-score frequencies at commonly used cut-offs and empirical base rates within the normative sample. Our secondary aim was to examine whether the choice of normative approach had implications for downstream clinical classification in the DDI cohort, particularly at the boundary between cognitively unimpaired (CU) status and MCI. To this end, we assessed three outcomes: concordance in classification of MCI versus CU, short-term diagnostic stability or change over approximately two years, and differences in established cerebrospinal fluid (CSF) biomarkers for Alzheimer’s Disease (AD) (Jack Jr. et al., 2024) across the resulting classification groups.

## Methods

### Cognitive tests

The DDI brief cognitive battery comprises the CERAD WLT learning and delayed recall subtests (Fillenbaum et al., 2008), TMT Parts A and B (Reitan & Wolfson, 1985), the FAS phonemic fluency test (Patterson, 2018), and the Silhouettes subtest from the Visual Object and Space Perception Battery (VOSP) (Warrington, 1991). Further details regarding administration and scoring are provided either in the original normative publications (Kirsebom et al., 2019; Espenes et al., 2020; Lorentzen et al., 2023) or in the case of VOSP silhouettes, the published test manual (Warrington, 1991).

### Normative samples

Our previous normative studies pooled healthy control data from three Nordic cohorts: the Norwegian DDI study, the Norwegian Trønderbrain study, and the Swedish Gothenburg Mild Cognitive Impairment study (G-MCI). DDI is a multicenter study primarily focused on early phases of dementia and Alzheimer’s disease across Norwegian health regions (Fladby et al., 2017). Trønderbrain is a Norwegian cohort based in the city of Trondheim established to study mild cognitive impairment, early Alzheimer’s disease, and healthy aging (Berge et al., 2016). The G-MCI study is a Swedish longitudinal research cohort focused on mild cognitive impairment, Alzheimer’s disease, and related neurodegenerative conditions (Wallin et al., 2016).

Across cohorts, healthy controls were selected to represent cognitively unimpaired adults within the age ranges available to each normative study. Although recruitment procedures varied somewhat between cohorts, controls were generally recruited from the community, from spouses or relatives of clinical participants, and through local recruitment initiatives. Broad inclusion criteria required absence of substantial cognitive symptoms, while major neurological, psychiatric, developmental, or severe somatic conditions likely to affect cognitive performance were exclusionary. More detailed descriptions of cohort design, recruitment, eligibility criteria, and assessment procedures have been published previously (Fladby et al., 2017; Berge et al., 2016; Wallin et al., 2016).

Because normative data availability differed across tests, sample size (n = 204–292) and age range (40 or 41 to 80 or 84 years) varied somewhat between measures. The DDI cohort contributed the majority of participants across studies (n = 168–170). To enable comparisons across the full neuropsychological battery, we identified participants who were represented in all previously published normative datasets (i.e., the intersection of the normative samples across studies; **Figure 1**). This resulted in a subsample of 131 participants with complete test data, all from the DDI cohort. This subsample was used to assess the expected frequency of low scores on each individual test at prespecified thresholds, as well as to calculate empirical base rates of obtaining *k* low scores across the battery. Further details regarding these analyses are provided in the Statistical analyses section.

**Figure 1.**
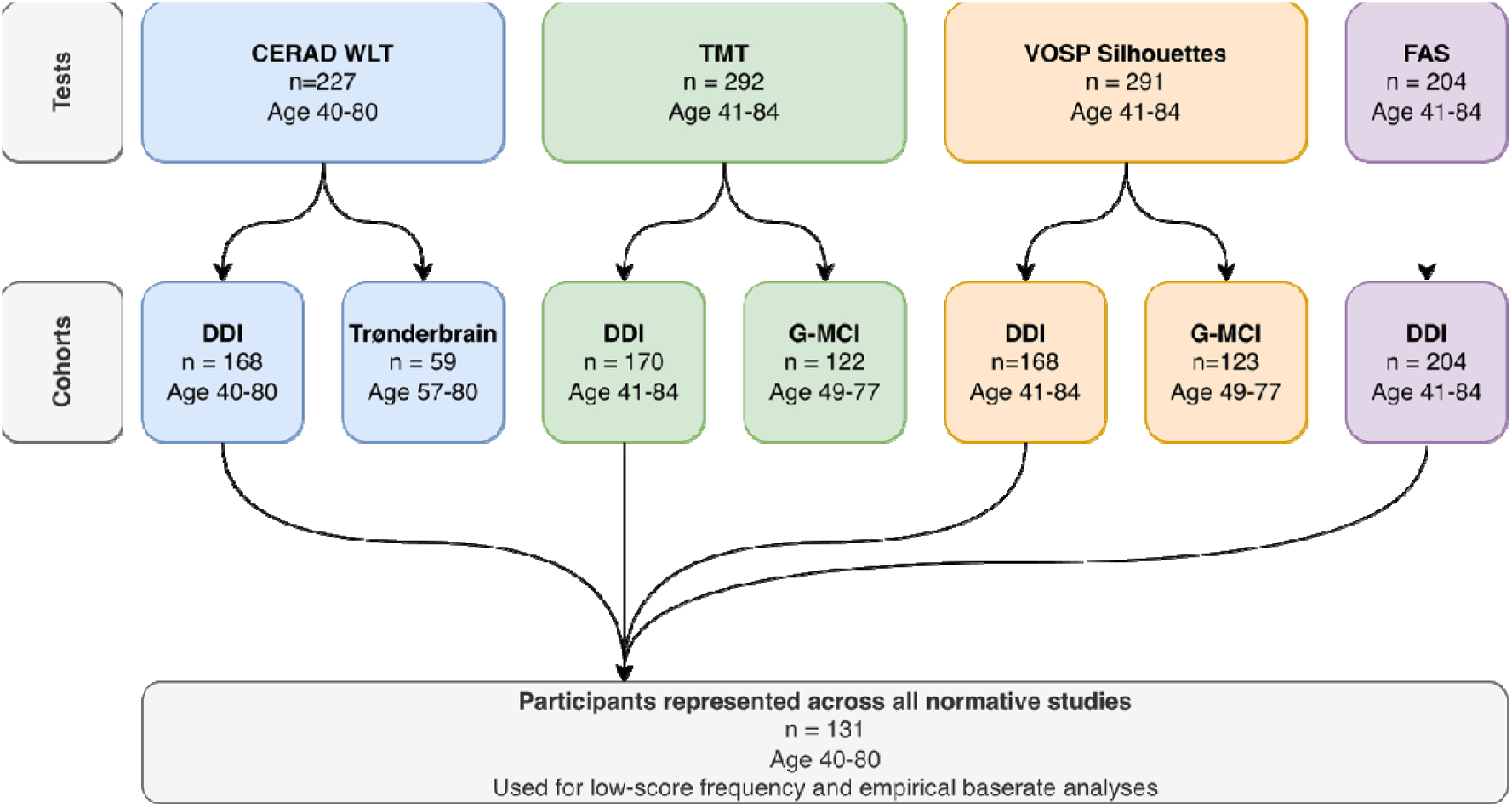
Overview of the control participants in each previous normative study, and participants represented across all studies. CERAD = Consortium to Establish a Registry for Alzheimer’s Disease, WLT = Word List Test, TMT = Trail Making Test, VOSP = Visual Object and Space Perception Battery. FAS = Letters F, A & S Phonemic Fluency Test. DDI = Dementia Disease Initiation, G-MCI = Gothenburg MCI.

### Previous LR-based norming procedures

In our previous normative studies, LR models were applied to standardized scaled scores rather than raw test scores. For most tests, including the CERAD word list subtests, Trail Making Test Parts A and B, and the VOSP Silhouettes test, raw scores were first transformed using cumulative percentile ranks and subsequently converted to standardized scaled scores (*M* = 10, *SD* = 3). These scaled scores were then used as outcome variables in linear regression models with demographic predictors. This transformation was introduced to improve adherence to LR assumptions. By contrast, for the FAS phonemic fluency test, raw scores were approximately normally distributed and no transformation was required; thus, LR was applied directly to raw scores. While this transformation approach resulted in acceptable model diagnostics, it also implies that multiple adjacent raw scores may map to the same scaled score, effectively reducing the resolution of the outcome variable.

### Clinical sample from the DDI cohort

The DDI cohort comprises individuals aged 40 to 80 years recruited from memory clinics and through advertisements in local news media across Norway. DDI focuses on the early preclinical (i.e. cognitively unimpaired, CU) and prodromal (MCI) stages of neurodegenerative disease and therefore does not include participants with dementia at baseline. Participants are reassessed at two-year intervals until study completion and may therefore progress to dementia during follow-up. See Fladby et al. (2017) for details.

For the present study, n = 643 participants from the DDI cohort with clinical symptoms (subjective and/or informant-based cognitive decline) and complete cognitive test data were included. One exception concerned the TMT, where failed performance (i.e. tests aborted because of cognitive inability to complete the task) was treated as the maximum value represented in the normative reference material (72 seconds for TMT-A and 167 seconds for TMT-B (Espenes et al., 2020)). Of these, 437 (68% of the baseline sample) had complete measurements of CSF Aβ (Amyloid beta) 42/40 ratio, phosphorylated tau at threonine 217 (p-tau217), total tau (t-tau), and neurofilament light chain (NfL) biomarkers (see CSF biomarkers section). However, please note that availability for CSF Aβ42/40 was greater (n=569, 88.5% of the baseline sample), and are included in the main descriptives. A total of 426 participants (66% of the baseline sample) had available cognitive follow-up data. For the diagnostic stability analysis, we included only the first available follow-up assessment as a measure of relatively short-term diagnostic stability and change, and restricted follow-up to visits occurring within 4 years of baseline. This excluded 14 cases, leaving 412 participants (64% of the original baseline sample). Mean follow-up time was 2.13 years (SD = 0.53; range = 0.30–3.87 years).

### Diagnostic criteria for Mild Cognitive Impairment and Dementia

In DDI, four of the measures in the brief battery are used as cognitive indicators in the assessment of MCI: verbal memory (CERAD delayed recall), executive functioning (TMT-B), language (FAS), and visuoperceptual ability (VOSP). This is consistent with the National Institute on Aging and Alzheimer’s Association (NIA-AA 2011) criteria for MCI (Albert et al., 2011), which define MCI as concern regarding a change in cognition with objective impairment in one or more cognitive domains, while independence in functional abilities is largely preserved. In the present study, MCI classifications were based on performance on these four indicators using either the original LR-based norms or the revised GAMLSS norms. For both approaches, objective cognitive impairment was operationalized as at least one test score with z ≤ −1.5, consistent with the operational criteria used in the DDI cohort (Fladby et al., 2017). Dementia diagnoses were established clinically according to the NIA-AA 2011 criteria for dementia due to Alzheimer’s disease (McKhann et al., 2011). These criteria require cognitive or behavioral symptoms that interfere with the ability to function at work or in usual activities and represent a decline from previous levels of functioning.

### CSF biomarkers

CSF biomarkers were selected from the DDI database to capture both AD-specific pathology and broader neurodegenerative injury. CSF Aβ42/40 ratio was included as a marker of amyloid plaque pathology (Hansson et al., 2019), while CSF p-tau217 was included as a tau phosphorylation marker closely associated with AD pathology and disease severity (Gonzalez-Ortiz et al., 2025; Janelidze et al., 2020). These markers align with the biomarker framework emphasized in the revised 2024 Alzheimer’s Association criteria, in which amyloid and phosphorylated tau biomarkers are central to biological AD classification (Jack Jr. et al., 2024). In addition, CSF t-tau and NfL were included as complementary markers of neuronal and neuroaxonal injury (Richter et al., 2025). Although these markers are less specific to AD than Aβ42/40 and p-tau217, they may provide additional information regarding neurodegenerative burden in clinically heterogeneous samples. CSF t-tau is elevated in AD but also in conditions involving acute (Franz et al., 2003), or rapid neuronal injury (Skillbäck et al., 2014), whereas NfL is widely regarded as a non-specific marker of neuroaxonal degeneration (Mielke et al., 2021; Yuan et al., 2017) and is associated with cerebrovascular disease (Meeker et al., 2022).

Lumbar punctures were performed according to the standardized BIOMARKAPD protocol, as previously described (Reijs et al., 2015). CSF Aβ1-42, Aβ1-40, NfL, and t-tau measurements were performed at Akershus University Hospital, Norway. Aβ1-42, Aβ1-40, and NfL were measured using the QuickPlex SQ120 platform (Meso Scale Discovery [MSD], MD, USA). Aβ1-42 and Aβ1-40 were analyzed in a multiplex format using the V-PLEX Aβ Peptide Panel 1 (6E10) kit (K15200E-1), whereas NfL was measured using the R-PLEX Human Neurofilament L Assay (K1517XR-2). The CSF Aβ42/40 ratio was calculated and amyloid positivity defined according to a previously established cohort-specific cut-off (Siafarikas et al., 2021). CSF concentrations of t-tau were measured using commercial enzyme-linked immunosorbent assays (INNOTEST, Fujirebio, Ghent, Belgium) based on monoclonal antibodies. CSF p-tau217 measurements were performed at the University of Gothenburg, Sweden, with one in 30 dilution factor according to the previously published method. (Gonzalez-Ortiz et al., 2024). Signal variations within and between analytical runs were assessed using 3 internal quality control samples at the beginning and the end of each run with a coefficient of variation below 15%.

### Statistical analyses

#### Descriptives

All analyses were conducted using RStudio (R version 4.3.2). Descriptive data for each normative study, including assessments of between-cohort differences and similarities in demographics and raw test scores, have been published previously. Accordingly, no new descriptive analyses were conducted for the present study. However, previously published data are summarized in the supplementary material. Descriptive statistics for the DDI clinical sample are presented for the baseline concordance sample, the longitudinal subsample, and the CSF biomarker subsample. Continuous variables are summarized as mean and standard deviation, and categorical variables as n (%).

### GAMLSS norming procedures

The GAMLSS framework (Stasinopoulos et al., 2017; Rigby et al., 2019) provides access to a wide range of probability distributions, allowing flexible modelling of outcome data that may deviate from the assumptions underlying standard regression approaches. To identify candidate distributions for each test, we used the model structures from the previously published LR regression-based norms as a starting point. For example, for the CERAD WLT subtests, age, sex, and years of education were initially specified as predictors of the location (μ) parameter. Candidate probability distributions were then evaluated using the chooseDist() function, which fits a predefined set of distributions to a specified model and ranks them according to the Generalized Akaike Information Criterion (GAI C). Two distribution families were considered: (1) binomial-type distributions for subtests with bounded integer score ranges from 0 to a fixed maximum value, such as the CERAD WLT (range 0 - 10) and VOSP Silhouettes tests (range 0 - 30), and (2) continuous distributions available in GAMLSS. For tests with a theoretical lower bound of 0 (that is, CERAD and VOSP measures), a small constant (0.001) was added prior to fitting continuous distributions. The best-fitting distribution for each test, according to GAIC, was then selected for further model building. Because most of the original LR normative models were fitted to transformed scaled scores rather than raw data, direct comparison of model fit statistics (e.g., AIC or BIC) with GAMLSS models is not appropriate. Instead, we compare the best-fitting GAMLSS distribution to a Gaussian GAMLSS model with the same predictor structure to isolate the effect of distributional assumptions while holding the predictor structure constant.

A stepwise model selection procedure was used to identify predictors for the location (μ), scale (σ), and when relevant also the shape parameters (ν for skewness, τ for kurtosis) within the selected distribution. Predictor entry was informed by the previously published norms. For example, for the CERAD tests, age was entered first, followed by sex and years of education (Kirsebom et al., 2019), whereas for FAS phonemic fluency, years of education was entered first, followed by age and sex (Lorentzen et al., 2023). For continuous predictors (age and years of education), penalized splines were also evaluated to assess potential non-linear associations. Predictors were retained only if they improved model fit according to GAIC and were statistically significant (p < 0.05). Model adequacy was assessed using diagnostic plots based on normalized quantile residuals and worm plots which were used to evaluate overall fit and detect localized deviations from the assumed distribution.

Because coefficients from GAMLSS models are not always directly interpretable in the same way as coefficients from LR, the Results section provides simple descriptive summaries of demographic influences for each test. For completeness, model statistics for the final normative GAMLSS models are provided in the supplementary material. For each final model, normative look-up tables were generated across the relevant ranges of age, sex, education, and raw test scores. For each covariate and raw-score combination, the fitted model was used to predict the distributional parameters and derive percentiles and corresponding z-scores. For tests where higher scores reflect better performance, we used the standard cumulative probability directly. For time-based tests, where higher scores reflect worse performance, we reversed the direction so that lower percentiles and more negative z-scores consistently indicate poorer performance. For discrete bounded outcomes, *mid-p* probabilities were used to avoid overly extreme percentile values at discrete score steps. Probabilities were constrained to avoid infinite z-scores at the distributional extremes.

The norms are presented in an online calculator (https://ddinorms.shinyapps.io/gamlss/), with optional look-up tables provided as supplementary material for use in research cohorts.

### Assessment of expected low-score frequencies, empirical base rates, and effects of LR transformations

In the subsample of 131 participants represented across all normative studies, we assessed the frequency of low scores on each individual test at thresholds of z ≤ −1 and z ≤ −1.5, corresponding to the 15.9th and 6.7th percentiles, respectively. These thresholds were chosen because they reflect commonly applied criteria for impaired test performance in the target population, particularly in the context of mild cognitive impairment due to neurodegenerative disease (Albert et al., 2011). For well-calibrated norms, the observed proportion of low scores on each individual test should approximate these theoretical percentile-based expectations (Crawford et al., 2009). To assess this, we used two-tailed proportion tests, where a significant result (p < 0.05) was interpreted as evidence of departure from the expected proportion. We also estimated empirical base rates for obtaining at least k low scores across tests at each threshold, with k corresponding to the number of individual tests in the battery (1 to 6).

### Assessment in clinical cohort

Concordance rates of MCI and CU were tabulated and assessed using Cohen’s kappa. We assessed potential differences in CSF biomarkers between discordant (e.g. MCI only with LR approach) and concordant classification groups (i.e. both CU or MCI) using linear regression models with log-transformed biomarkers as outcomes and age and sex as covariates. The Benjamin-Hochberg (BH) approach was used to control the false discovery rate within each comparison (four tests, corresponding to each biomarker). Lastly, short-term diagnostic change was tabulated for each normative approach and summarized as n (%). Transitions considered likely to differ between approaches were formally compared using McNemar’s test.

### Ethics

The DDI, TrønderBrain, and Gothenburg MCI studies were approved by their respective regional or local research ethics committees. All participants provided written informed consent prior to participation, including information regarding the right to withdraw and the potential risks and benefits of participation. All datasets used in the present study were de-identified prior to analysis, with participants represented only by study-specific identification codes; linkage keys and directly identifying information were not included in the analytic datasets. All study procedures were conducted in accordance with the ethical principles of the Declaration of Helsinki (1964, revised 2013) and applicable national regulations, including the Norwegian Health Research Act where relevant.

## Results

### GAMLSS norms

Detailed model structures and coefficients are provided in Supplementary Table 2, with GAIC-based model comparisons in Supplementary Table 3. Worm plots for the selected GAMLSS and corresponding Gaussian models are presented side-by-side in Supplementary Figure 1.

For both CERAD WLT learning (Figure 2A) and delayed recall (Figure 2B), the beta-binomial (BB) distribution provided the best fit. The final models retained age, sex, and years of education as predictors of the location parameter, with the scale parameter held constant. For both subtests, higher age was associated with lower expected performance (both p < 0.001), whereas female sex (both p ≤ 0.01) and more years of education (both p ≤ 0.05) were associated with better performance.

**Figure 2.**
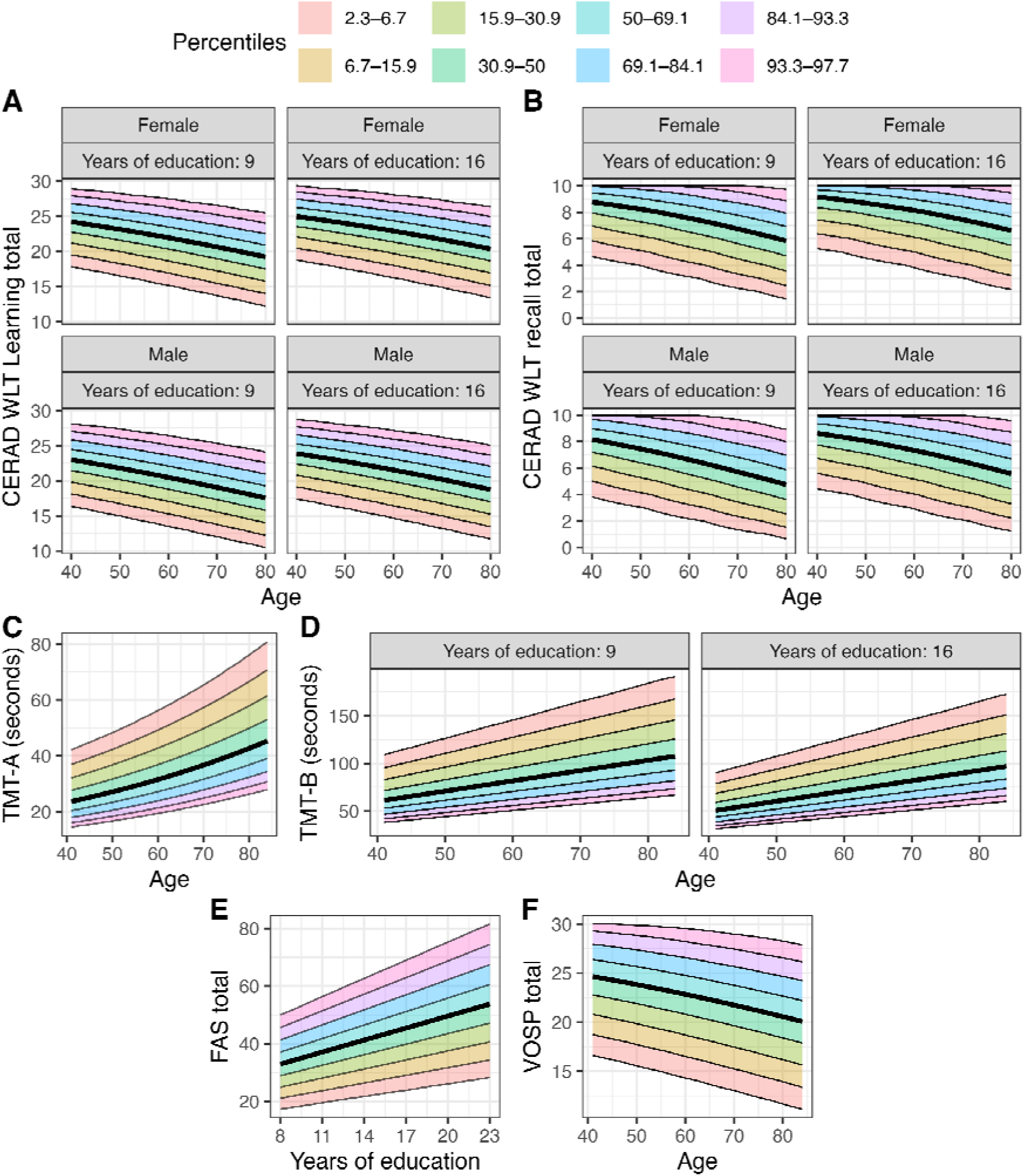
Illustration of the final Generalized Additive Models for Location, Scale and Shape (GAMLSS) normative models for the Consortium to Establish a Registry for Alzheimer’s Disease (CERAD) Word List Test (WLT) Learning (**A**) and Delayed Recall (**B**), Trail Making Test Parts A (**C)** and B (**D**), FAS phonemic fluency (**E**), and the Silhouettes subtest from the Visual Object and Space Perception Battery (VOSP) (**F**). For the CERAD WLT subtests and TMT-B, years of education were included as linear predictors in the models and are shown here at lower (9 years) and higher (16 years) education levels for illustration.

For TMT-A and TMT-B, the Box-Cox Power Exponential original (BCPEo) and Box-Cox Power Exponential (BCPE) distributions provided the best fit, respectively. For both tests, scale and shape parameters were modeled as constants. The final TMT-A model (Figure 2C) retained age as a predictor of completion time, with higher age associated with longer completion time (p < 0.001). The final TMT-B model (Figure 2D) retained both age and years of education, with higher age associated with longer completion time (p < 0.001) and more years of education associated with shorter completion time (p < 0.001).

For FAS phonemic fluency (Figure 2E), the Box-Cox Cole and Green (BCCG) distribution provided the best fit, although only marginally better than the normal distribution (GAIC = 1535 vs 1536). The final model retained years of education as a predictor of the location parameter, with higher education associated with better phonemic fluency performance (p < 0.001). Scale and skewness were modeled as constants.

For VOSP Silhouettes (Figure 2F), the BB distribution provided the best fit. The final model retained age as a predictor of the location parameter, with higher age associated with lower performance (p < 0.001). The scale parameter was held constant.

### Single-Test Low-Score Frequencies and Empirical Base Rates

Compared with the previous LR norms, the GAMLSS norms yielded lower frequencies of low scores across the battery (Figure 3A). At the 16th-percentile cut-off, the proportion with at least one low score was 53.4% under GAMLSS versus 63.4% under LR. At the 6.7th-percentile cut-off, the corresponding figures were 30.5% versus 40.5%. Differences were smaller at higher values of k, but the same overall pattern remained.

**Figure 3.**
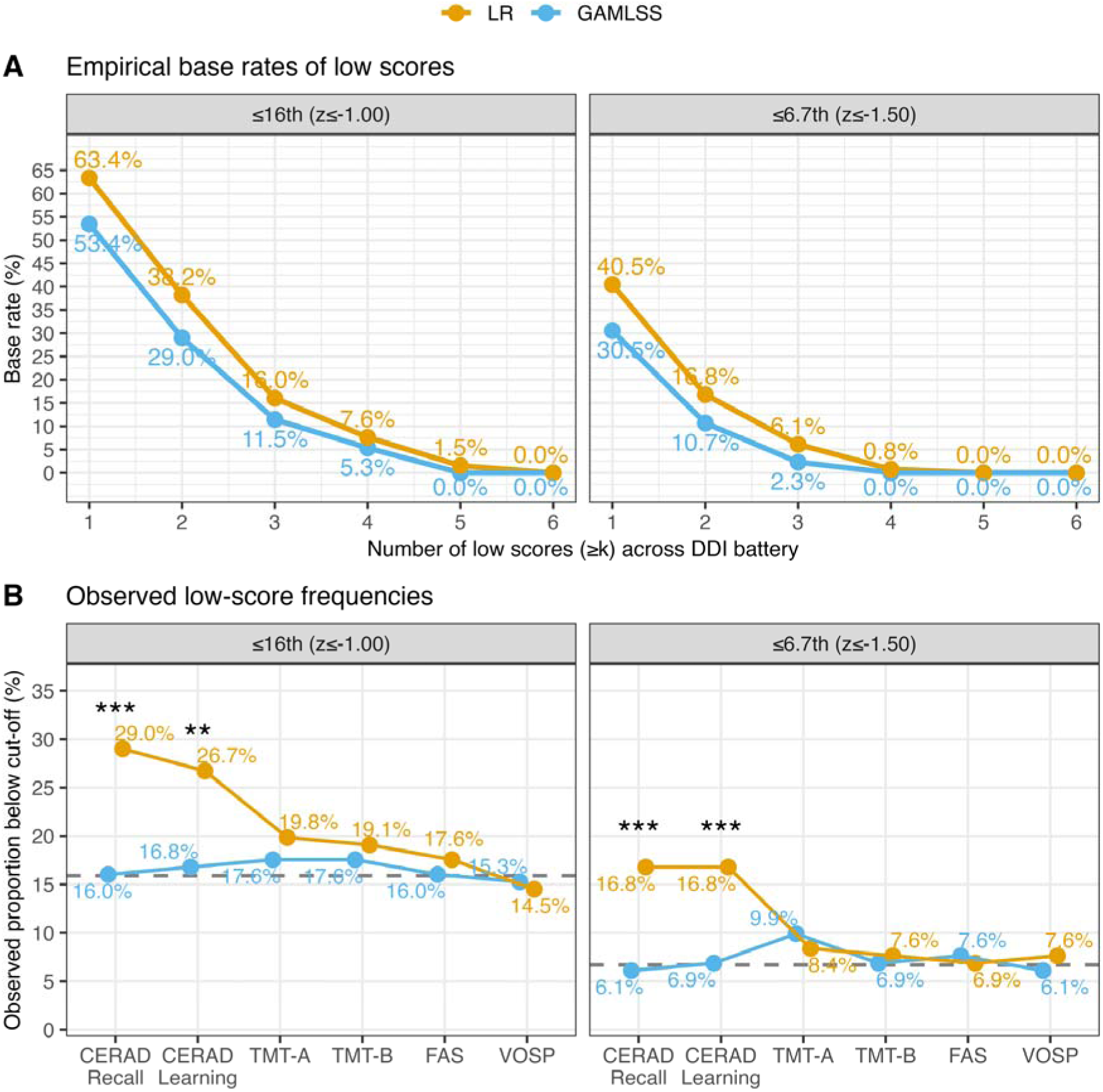
Panel **A** shows empirical base rates for obtaining at least k low scores at the 15.9th (z ≤ −1) and 6.7th (z ≤ −1.5) percentile thresholds in the subsample represented across all normative studies (n = 131). Panel **B** shows the corresponding low-score frequencies for each individual test in the same subsample. CERAD = Consortium to Establish a Registry for Alzheimer’s Disease, TMT = Trail Making test, FAS = FAS phonemic fluency, VOSP = the Visual Object and Space Perception battery silhouettes test, DDI = Dementia Disease Initiation, LR = Linear Regression, GAMLSS = Generalized Additive Models for Location, Scale and Shape.

Analyses at the single-test level indicated that this discrepancy was driven mainly by CERAD learning and delayed recall, for which the previous LR norms classified substantially more individuals than expected below both conventional cut-offs than the GAMLSS norms (Figure 3B). By contrast, no significant differences between normative methods were observed for TMT-B, FAS phonemic fluency, or VOSP Silhouettes. This pattern prompted us to compare observed and expected proportions for the CERAD subtests in the original normative sample (n = 227). In this sample, the original LR norms showed no significant misalignment at the z ≤ −1 threshold (15.9th percentile; learning: 18.1% observed, p = 0.365; delayed recall: 15.8% observed, p = 0.975). At the z ≤ −1.5 threshold (6.7th percentile), learning again closely matched expectation (6.7% observed, p = 0.965), whereas delayed recall showed a numerically higher observed proportion of low scores (9.6%), although this did not reach statistical significance (p = 0.072).

### Scaled-score transformation introduces stepwise normative z-score profiles

Across tests, the original LR-based norms exhibited a stepwise relationship between raw scores and z-scores, reflecting the discretization introduced by the scaled-score transformation (Figure 4). This resulted in plateaus of identical z-scores across adjacent raw scores, followed by abrupt transitions at category boundaries. In contrast, GAMLSS-derived z-scores varied smoothly across the full score range. For bounded measures where very low raw scores were rare (e.g., CERAD learning and VOSP Silhouettes), the GAMLSS beta-binomial models yielded more extreme z-scores at the lower end of the distribution, whereas the LR scaled-score approach truncated these values at a higher threshold. For FAS phonemic fluency, where no transformation was applied, the two approaches showed near-identical continuous relationships.

**Figure 4.**
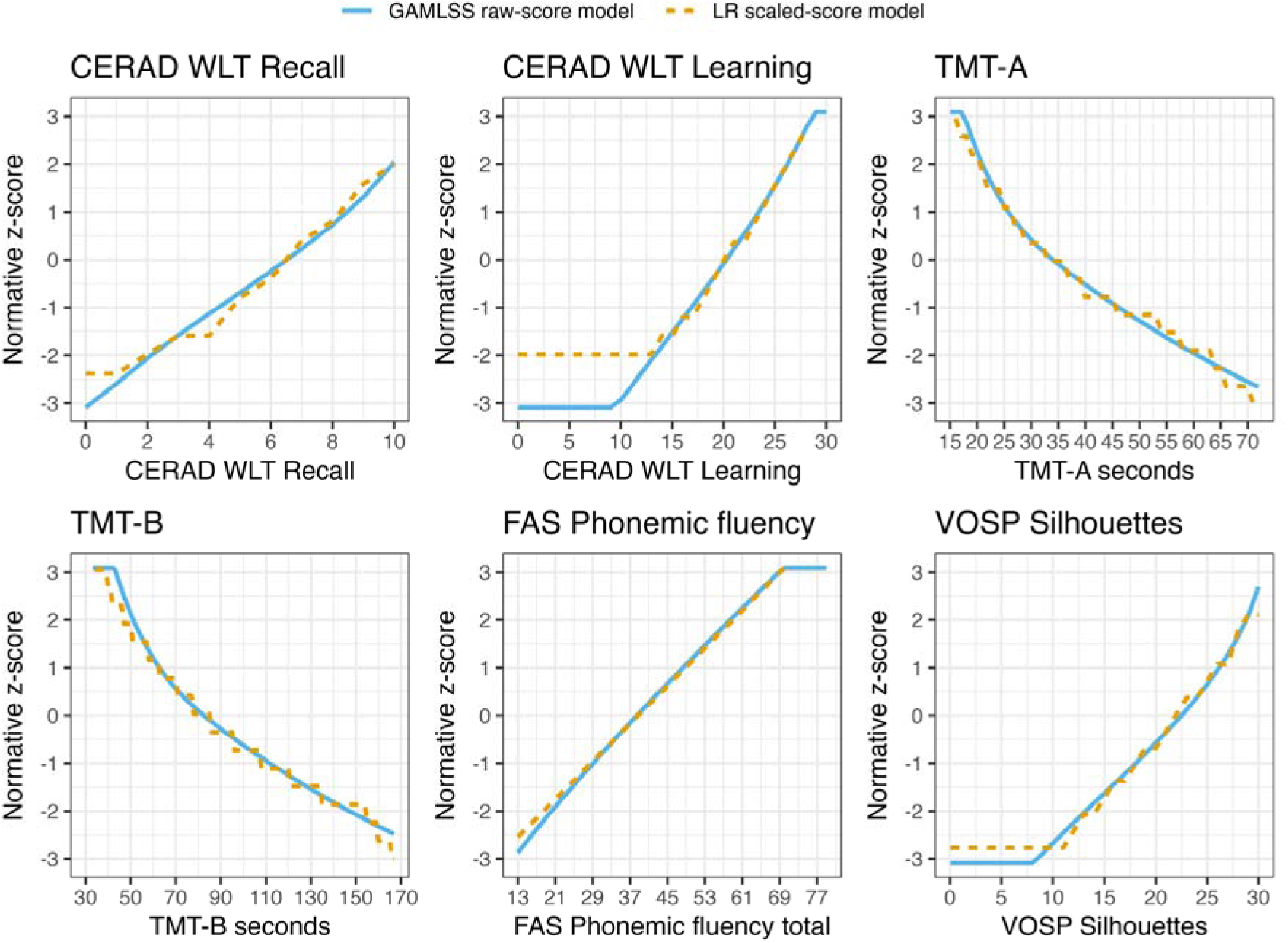
Relationship between raw scores and normative z-scores for the Generalized Additive Models for Location, Scale and Shape (GAMLSS) raw-score models and the previous linear regression (LR) norms, illustrated for a hypothetical 65-year-old male with 12 years of education. For the Consortium to Establish a Registry for Alzheimer’s Disease Word List Test (CERAD WLT), Trail Making Test (TMT), and the Visual Object and Space Perception Battery (VOSP) Silhouettes test, the previous LR norms were based on transformed scaled-score outcomes. By contrast, FAS phonemic fluency was modeled directly using raw scores. The stepwise LR profiles illustrate the discretization introduced by the scaled-score transformation.

### Concordance of diagnoses between normative approaches

Please see Table 1 for descriptive statistics of the DDI cross-sectional concordance sample. Concordance between the two normative approaches was high (Cohen’s κ = 0.913, p < 0.001, See figure 5A). Of the full sample (n = 643), 246 (38.3%) were concordantly classified as CU and 370 (57.5%) as MCI, leaving 27 cases (4.2%) classified discordantly. Only four cases were classified as MCI by the GAMLSS norms but not by the LR norms, reflecting isolated borderline impairments on FAS phonemic fluency (n = 2), TMT-B (n = 1), and VOSP Silhouettes (n = 1). By contrast, the remaining 23 cases were classified as CU under the GAMLSS norms but as MCI under the LR norms. Of these, 22 (95.7%) were driven by CERAD delayed recall scores that fell below the LR threshold but not the GAMLSS threshold, whereas the remaining case reflected a borderline FAS phonemic fluency score.

**Figure 5.**
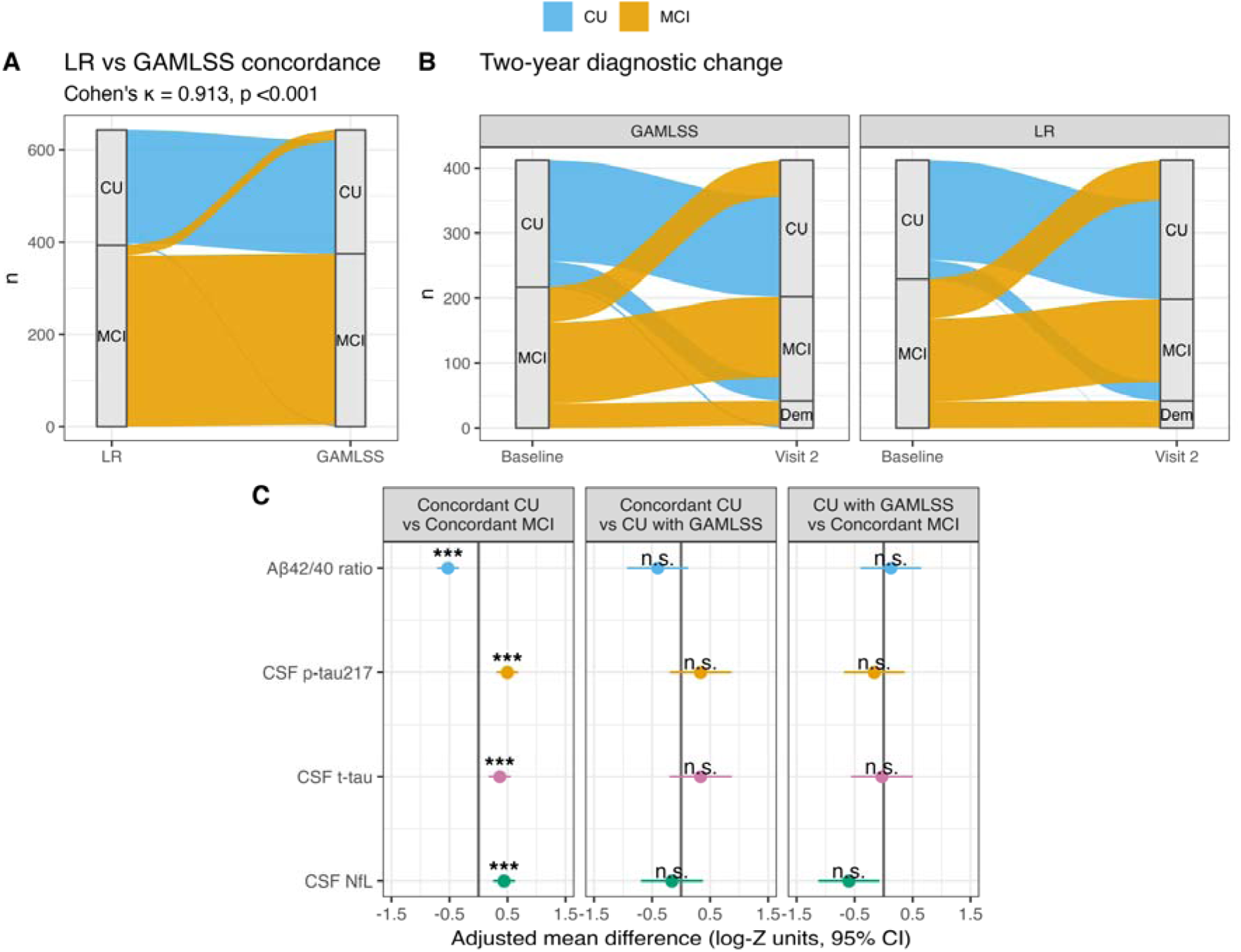
**Panel A** shows the concordance rates of baseline Cognitively Unimpaired (CU) and Mild Cognitive Impairment (MCI) classifications using Generalized Additive Models for Location Scale and Shape (GAMLSS) or Ordinary Least Squares linear regression (LR) within the Dementia Disease Initiation clinical sample. **Panel B** shows short-term diagnostic stability and change of diagnoses using these different normative approaches. **Panel C** shows the comparisons of cerebrospinal fluid (CSF) Aβ (amyloid beta) 42/40 ratio, phosphorylated tau at threonine 217 (p-tau217), total-tau (t-tau) and Neurofilament Light Chain (NfL) between concordant and discordant CU/MCI groups. Biomarkers were log-transformed and standardized before analysis. Mean differences (95% confidence intervals) were adjusted for age and sex. P-values were adjusted using the Benjamini–Hochberg procedure.

**Table 1.** Demographic and cognitive characteristics by concordance group in the Dementia Disease Initiation clinical sample.

| Variable | Total | Concordant<br>CU | Concordant<br>MCI | CU<br>with GAMLSS | MCI<br>with GAMLSS |
| --- | --- | --- | --- | --- | --- |
| N | 643 (100%) | 246 (38.3%) | 370 (57.5%) | 23 (3.6%) | 4 (0.6%) |
| **Female,**<br>n (%) | 321 (49.9%) | 131 (53.3%) | 171 (46.2%) | 17 (73.9%) | 2 (50.0%) |
| **Aβ42/40 positive,**<br>n (%) [missing] | 256 (45.0%)<br>[74] | 76 (34.4%)<br>[25] | 168 (51.7%)<br>[45] | 11 (52.4%)<br>[2] | 1 (50.0%)<br>[2] |
| **Age** | 64.3 (8.8) | 62.9 (8.8) | 65.2 (8.8) | 63.4 (7.9) | 71.2 (7.5) |
| **Years of education** | 13.7 (3.0) | 13.9 (2.9) | 13.6 (3.1) | 14.0 (2.2) | 11.5 (3.7) |
| **CERAD WLT recall raw,** | 5.0 (2.8) | 7.1 (1.8) | 3.6 (2.6) | 4.8 (1.3) | 5.8 (1.7) |
| **CERAD WLT recall GAMLSS z** | -1.0 (1.2) | -0.1 (0.8) | -1.5 (1.1) | -1.2 (0.5) | -0.3 (0.7) |
| **CERAD WLT recall LR z** | -0.9 (1.2) | -0.1 (0.9) | -1.5 (1.1) | -1.5 (0.5) | -0.5 (0.9) |
| **CERAD WLT learning**<br>**GAMLSS z** | -0.9 (1.3) | -0.1 (1.0) | -1.4 (1.1) | -1.1 (1.0) | -1.3 (1.2) |
| **CERAD WLT learning LR z** | -0.8 (1.1) | -0.1 (1.0) | -1.3 (1.0) | -1.1 (0.9) | -1.0 (1.0) |
| **TMT-A seconds** | 45.0 (26.8) | 34.6 (12.4) | 52.4 (31.9) | 37.7 (10.8) | 42.5 (7.7) |
| **TMT-A GAMLSS z** | -0.5 (1.3) | 0.1 (1.1) | -0.9 (1.3) | -0.3 (0.8) | -0.4 (0.5) |
| **TMT-A LR z** | -0.5 (1.2) | 0.1 (1.1) | -0.9 (1.2) | -0.3 (0.7) | -0.3 (0.5) |
| **TMT-B seconds** | 123.8 (76.5) | 78.4 (22.6) | 156.8 (85.1) | 80.6 (16.5) | 113.2 (27.3) |
| **TMT-B GAMLSS z** | -0.9 (1.5) | 0.1 (1.0) | -1.6 (1.3) | -0.1 (0.5) | -0.7 (0.6) |
| **TMT-B LR z** | -0.8 (1.4) | 0.1 (1.0) | -1.5 (1.3) | -0.1 (0.5) | -0.6 (0.5) |
| **FAS raw** | 37.2 (12.8) | 43.3 (10.3) | 33.1 (12.9) | 39.2 (7.8) | 28.8 (11.6) |
| **FAS GAMLSS z** | -0.4 (1.2) | 0.2 (0.9) | -0.8 (1.2) | -0.2 (0.7) | -1.1 (0.8) |
| **FAS LR z** | -0.4 (1.2) | 0.2 (1.0) | -0.8 (1.2) | -0.2 (0.7) | -0.9 (0.8) |
| **VOSP Silhouettes raw** | 21.0 (4.5) | 23.2 (3.5) | 19.5 (4.6) | 22.0 (3.5) | 19.8 (3.9) |
| **VOSP Silhouettes GAMLSS z** | -0.3 (1.1) | 0.2 (1.0) | -0.6 (1.0) | -0.1 (0.9) | -0.4 (0.9) |
| **VOSP Silhouettes LR z** | -0.3 (1.1) | 0.2 (0.9) | -0.6 (1.1) | -0.1 (0.9) | -0.4 (1.0) |

Notes. GAMLSS = Generalized Additive Models for Location, Scale and Shape; LR = linear regression; CERAD WLT = Consortium to Establish a Registry for Alzheimer's Disease Word List Test; TMT = Trail Making Test; FAS = phonemic fluency (letters F, A, and S); VOSP = Visual Object and Space Perception Battery. Values are mean (SD) unless otherwise specified. Female and Aβ42/40 positivity are shown as n (%).

### Two-year diagnostic change

Please see Table 2 for descriptive statistics regarding the longitudinal subsample. Broadly speaking, the two normative approaches yielded very similar 2-year transition patterns across the 412 cases with longitudinal data. For GAMLSS versus LR, respectively, 155 (37.6%) versus 153 (37.1%) remained CU, 124 (30.1%) versus 127 (30.8%) remained MCI, 36 (8.7%) versus 29 (7.0%) progressed from CU to MCI, 55 (13.3%) versus 61 (14.8%) reverted from MCI to CU, 38 (9.2%) versus 41 (10.0%) progressed from MCI to dementia, and 4 (1.0%) versus 1 (0.2%) progressed from CU to dementia. The only notable differences were small shifts in transition rates, with LR norms yielding 1.5% more MCI-to-CU reversions and GAMLSS norms yielding 1.7% more CU-to-MCI conversions. However, neither difference reached statistical significance (McNemar’s χ²(1) = 1.25, p = 0.263; χ²(1) = 2.11, p = 0.146). See figure 5B.

**Table 2.**
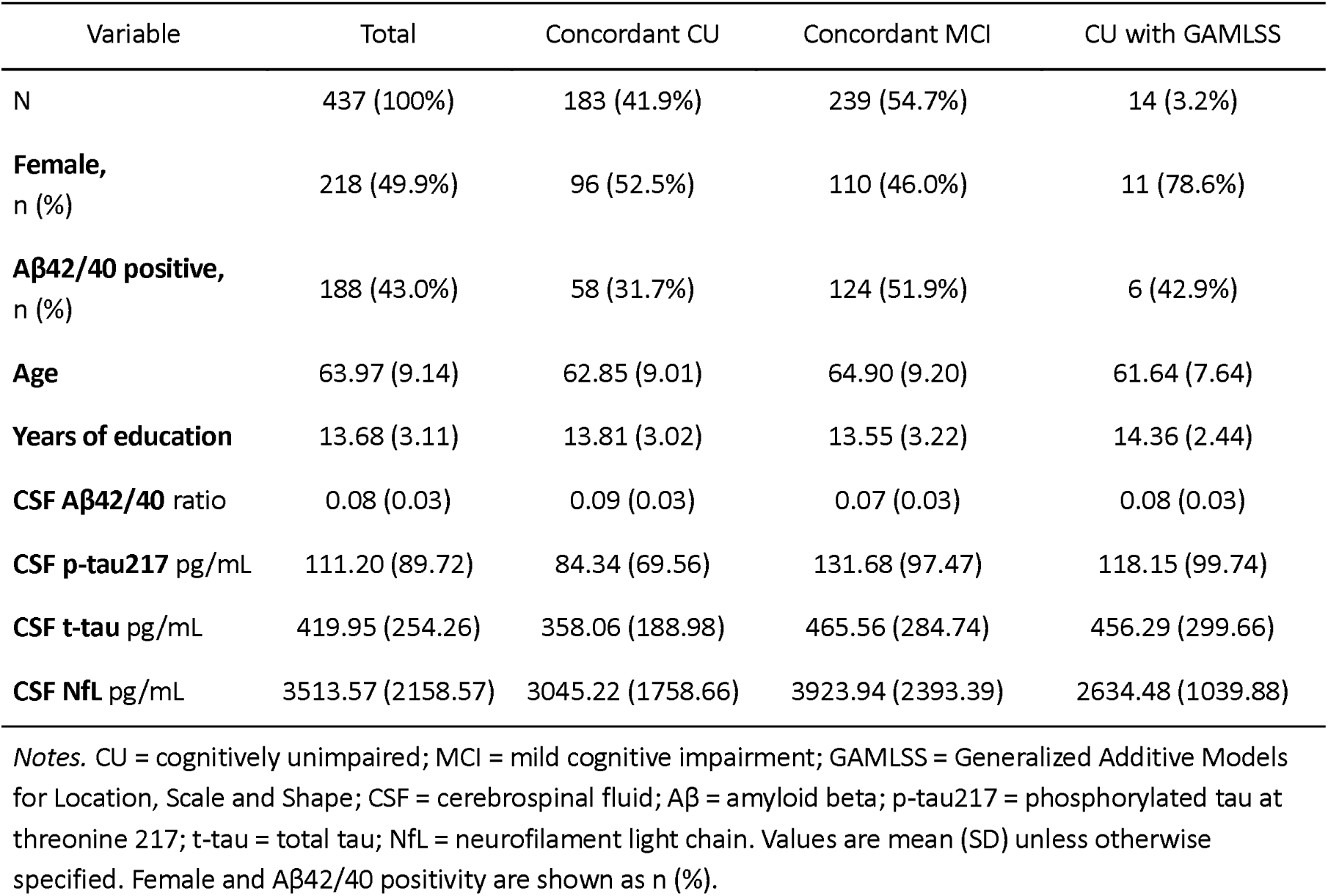
Demographic and CSF biomarker characteristics by concordance group.

### Comparison of biomarkers of neurodegenerative disease

Please see Table 3 for descriptive statistics regarding the CSF subsample.

**Table 3.** Demographic characteristics by short-term diagnostic transition group.

|  | Variable | Total | Stable CU | Stable MCI | CU to MCI | MCI to CU | CU to Dementia | MCI to Dementia |
| --- | --- | --- | --- | --- | --- | --- | --- | --- |
| GAMLSS Transition groups | **N, %** | 412 (100%) | 155 (37.6%) | 124 (30.1%) | 36 (8.7%) | 55 (13.3%) | 4 (1.0%) | 38 (9.2%) |
|  | **Female, n (%)** | 218 (52.9%) | 87 (56.1%) | 53 (42.7%) | 22 (61.1%) | 38 (69.1%) | 2 (50.0%) | 16 (42.1%) |
|  | **Aβ42/40 positive, n (%)**<br>[Missing] | 164 (43.4%)<br>[34] | 45 (31.5%)<br>[12] | 52 (46.4%)<br>[12] | 15 (41.7%) | 13 (28.9%)<br>[10] | 4 (100.0%) | 35 (92.1%) |
|  | **Baseline age** | 64.3 (9.0) | 62.7 (8.8) | 64.7 (8.8) | 66.7 (8.3) | 62.0 (10.0) | 71.8 (4.5) | 69.2 (7.3) |
|  | **Years of education** | 13.7 (3.1) | 14.0 (2.9) | 13.7 (3.4) | 13.4 (2.5) | 13.7 (3.2) | 12.2 (2.1) | 13.1 (3.2) |
|  | **Years since baseline\*** | 2.1 (0.5) | 2.2 (0.5) | 2.1 (0.6) | 2.3 (0.3) | 2.0 (0.6) | 1.8 (0.2) | 1.9 (0.6) |
| LR Transition groups | **N, %** | ** | 153 (37.1%) | 127 (30.8%) | 29 (7%) | 61 (14.8%) | 1 (0.2%) | 41 (10%) |
|  | **Female, n (%)** | ** | 84 (54.9%) | 59 (46.5%) | 17 (58.6%) | 40 (65.6%) | 0 (0.0%) | 18 (43.9%) |
|  | **Aβ42/40 positive, n (%)**<br>[Missing] | ** | 43 (30.3%)<br>[11] | 54 (46.6%)<br>[11] | 12 (42.9%)<br>[1] | 16 (32.0%)<br>[11] | 1 (100.0%) | 38 (92.7%) |
|  | **Baseline age** | ** | 63.6 (8.9) | 64.6 (8.8) | 64.4 (9.2) | 61.7 (9.6) | 77.0 | 69.2 (7.1) |
|  | **Years of education** | ** | 13.8 (2.9) | 13.9 (3.3) | 13.5 (2.9) | 13.7 (3.1) | 10.0 | 13.1 (3.1) |
|  | **Years since baseline\*** | ** | 2.2 (0.5) | 2.1 (0.5) | 2.2 (0.3) | 2.1 (0.6) | 2.0 | 1.9 (0.6) |

Notes. Aβ = Amyloid beta; CU = cognitively unimpaired; MCI = mild cognitive impairment; GAMLSS = Generalized Additive Models for Location, Scale and Shape; LR = linear regression. Values are mean (SD) unless otherwise specified. Total N per group, female sex and Aβ42/40 ratio positivity are shown as n (%). Values in square brackets indicate number of missing observations.

CSF biomarkers were available for 437 of 643 cases (68%). This resulted in reduced numbers in the concordance groups, particularly for the discordant CU/MCI cases of interest, where 14 of 23 had complete data for the chosen biomarkers. Concordant MCI cases were associated with lower CSF Aβ42/40 ratios than concordant CU cases (p < 0.001), as well as higher p-tau217 (p < 0.001), higher t-tau p < 0.001) and higher NfL (p < 0.001). The discordant cases classified as CU by GAMLSS but MCI by LR (denoted as “CU with GAMLSS” in Figure 5C) did not differ significantly from either concordant CU or concordant MCI cases on any biomarker. Only 1 of 4 of the discordant cases classified as MCI by GAMLSS but CU by LR had available CSF data and was therefore dropped from the analysis.

## Discussion

In this study, we revisited our previously published LR regression-based norms for the DDI brief battery using the same normative samples, but within a GAMLSS framework. For most tests, the two approaches produced comparable rates of low scores at commonly used cut-offs. However, the CERAD WLT learning and recall tests produced large discrepancies in observed versus expected proportions at the 15.9th and 6.7th percentiles within the normative subsample. Moreover, the scaled-score transformation in the original LR models introduced a stepwise relationship between raw scores and z-scores, with plateaus across adjacent scores and abrupt transitions at category boundaries. In contrast, GAMLSS-derived z-scores showed smooth and continuous relationships with raw performance across the full score range. Despite these differences, we found no clear downstream clinical advantage of either normative approach in the clinical sample. Concordance between CU and MCI classification was high, with only 4.2% classified discordantly. These discrepancies were driven largely by stricter LR normative scores for CERAD delayed recall. Similarly, short-term diagnostic stability and change were broadly comparable. In the CSF biomarker analyses, concordant MCI cases showed an AD-related profile (lower Aβ42/40 ratio and higher p-tau217), including elevated markers of neurodegeneration (higher t-tau and NfL), compared with concordant CU cases. The discordant group did not differ significantly from either group across any biomarker.

The most noteworthy findings involved the CERAD WLT learning and delayed recall subtests, which showed higher-than-expected proportions of low scores in the normative subsample. This pattern was most pronounced for delayed recall and was not observed for the corresponding GAMLSS norms, which closely matched expected proportions. Although less evident in the full normative sample, delayed recall still showed a modest elevation at the 6.7th percentile under the original LR norms, suggesting that subsample characteristics alone do not fully explain the findings. Consistent with this, stricter LR-based delayed recall norms were the primary driver of diagnostic disagreement in the clinical sample. A likely explanation is that CERAD delayed recall exhibits mild skewness in cognitively healthy middle-aged individuals (Kirsebom et al., 2019). The test’s restricted 10-word range

(Fillenbaum et al., 2008) may limit resolution in higher-functioning individuals. Under the LR framework, this was further discretized through scaled-score transformation, grouping adjacent raw scores into broader categories and producing stepwise patterns in normative z-scores, particularly at the distribution tails. Consequently, small differences in raw performance may have resulted in larger shifts in normative classification around impairment thresholds. For CERAD learning, low raw scores were rare. Combined with the scaled-score transformation, this compressed the lower end of the distribution, mapping limited variability into coarse categories. As a result, small differences near the lower bound may have led to larger shifts in classification, contributing to inflated low-score frequencies. In contrast, GAMLSS beta-binomial models directly modeled the bounded raw-score distributions and preserved a continuous relationship between raw scores and z-scores, resulting in closer alignment with expected proportions.

The key advantage of GAMLSS for continuous norming is its ability to model multiple aspects of the outcome distribution, allowing demographic predictors to influence not only expected performance, but also variability and distributional shape when supported by the data (Stasinopoulos et al., 2017; Rigby et al., 2019). This enabled direct modeling of raw scores using distributions that better reflected observed data characteristics, rather than relying on transformations to meet linear regression assumptions. Accordingly, improved model fit over the Gaussian GAMLSS equivalent was observed for all tests, and the selected distributions were broadly consistent with recent GAMLSS-based neuropsychological norming studies. In particular, bounded cognitive outcomes favored beta-binomial distributions and timed measures such as TMT-A/B favored Box-Cox power exponential distributions (Rubio-Guerra et al., 2025; Grigorova et al., 2025; Sachs et al., 2022). Moreover, the FAS phonemic fluency test showed only negligible improvement of the Box-Cox Cole and Green distribution over the Gaussian model, reflecting mild skewness and an approximately normal distribution in practical terms. This was also consistent with previous LR-based residual diagnostics (Lorentzen et al., 2023), and with recently developed GAMLSS norms for this measure (Rubio-Guerra et al., 2025). Nevertheless, the original LR norms were robust in the clinical analyses when applying predefined z-score cut-offs for MCI classification (Fladby et al., 2017; Albert et al., 2011). These findings suggest that conventional LR norms may remain robust when transformed outcomes are well calibrated and applied within a similar population.

However, the modest downstream differences may also reflect that demographic predictors influenced only the location parameter in the final GAMLSS models and were identical across approaches. GAMLSS may offer greater advantages in settings where demographic variables additionally influence variability or distributional shape (Timmerman et al., 2021).

Most discordant cases were classified as CU under GAMLSS but as MCI under LR norms and were largely driven by small differences in CERAD delayed recall around the z ≤ −1.5 threshold. These cases may represent borderline amnestic impairment but may also include false-positive classifications arising from reliance on a single low test score. This highlights a broader limitation of brief neuropsychological batteries, where the likelihood of obtaining at least one low score increases with the number of tests administered (Binder et al., 2009; Brooks et al., 2013). For example, at the 6.7th percentile threshold, the probability of at least one low score across six independent tests is approximately 34% [1 − (1 − 0.067)^6], closely aligning with the observed base rate in the GAMLSS norms (30.5%). Thus, one low score is not uncommon at the battery level, even among cognitively healthy individuals, consistent with previous empirical studies (Palmer et al., 1998; Mistridis et al., 2015; Holdnack et al., 2017).

This issue is directly relevant to MCI classification. Criteria that allow a single low score to define impairment are inherently sensitive to borderline test performance and may increase false-positive classifications. The reversion findings may in part be consistent with this interpretation. MCI-to-CU reversion is common in longitudinal studies and may reflect a mixture of false-positive baseline classification, practice effects, fluctuating cognition, and true recovery (Chung et al., 2019). Importantly, studies comparing conventional and actuarial/neuropsychological criteria have found that more comprehensive criteria can reduce reversion and identify groups with stronger biomarker and progression profiles (Wong et al., 2018; Jak et al., 2009). Alternative actuarial approaches, in particular the Jak/Bondi criteria (Jak et al., 2009; Bondi et al., 2014; Bondi et al., 2008), attempt to reduce this problem by requiring more consistent evidence of impairment, defined as at least two scores ≤ −1 SD within the same cognitive domain. Although this approach uses a less extreme single-test threshold, the requirement of multiple low scores within a domain lowers the likelihood that classification is driven by isolated low performance or normal measurement variability.

At the same time, the CSF biomarker findings suggest that at least some discordant cases may reflect biologically relevant borderline impairment rather than purely expected normative variability. Although the discordant group did not differ significantly from concordant CU cases on any biomarker, the overall pattern for CSF Aβ42/40 ratio, p-tau217, and t-tau was directionally consistent with a possible enrichment of AD-related pathology (Jack Jr. et al., 2024). Notably, 52.40% (11/21) of discordant CERAD delayed recall cases with available CSF Aβ42/40 data were amyloid-positive based on our cut-off (Siafarikas et al., 2021). Although CSF NfL levels were numerically, but not significantly, lower in the discordant group than in concordant MCI cases, this pattern may nevertheless be of interest. Given that CSF NfL is regarded as a non-specific marker of neuroaxonal degeneration (Mielke et al., 2021; Yuan et al., 2017) and has been linked to disease severity across neurodegenerative disorders (Delaby et al., 2020), the overall biomarker pattern may suggest a heterogeneous group comprising individuals with very early or borderline impairment alongside cases reflecting expected variability or potential false-positive classifications. However, amyloid-negative cases with memory impairment may also reflect non-AD etiologies, such as cerebrovascular disease (Rundek et al., 2022) or Lewy body pathology (Hemminghyth et al., 2020). As the present study was not designed to assess alternative causes, this interpretation remains speculative.

This study has some limitations. The normative subsample used to compare approaches was substantially smaller than the full normative datasets, and empirical base rates should therefore be considered approximations rather than precise estimates for this test battery. Nevertheless, this allowed assessment of the robustness of the GAMLSS norms, particularly for the CERAD subtests. Similarly, the reduced sample with available CSF biomarkers limits conclusions regarding the discordant cases. Finally, analyses were restricted to the first available follow-up to focus on relatively short-term diagnostic stability and change. While this allowed assessment of early instability in CU/MCI classification under the LR and GAMLSS norms, longer-term progression was not examined.

In conclusion, GAMLSS provided a more flexible and faithful representation of neuropsychological test score distributions, particularly for bounded and non-normal outcomes. Despite these advantages, clinical implications were modest, with high concordance in CU/MCI classification, similar short-term diagnostic stability and change, and largely comparable biomarker profiles. Differences between approaches were infrequent and primarily driven by CERAD delayed recall near diagnostic thresholds, suggesting that well-calibrated LR norms remain robust in clinical practice. Future work should evaluate GAMLSS in more complex neuropsychological settings, including tests with restricted ranges or where demographic factors influence variability or distributional shape, to better define when these modeling advantages translate into clinically meaningful improvements.

## Data Availability

Due to ethical restrictions, the data generated in the present study are not freely available for sharing.

## Declaration of generative AI use

ChatGPT versions 5.2 – 5.5 was used during manuscript preparation to support language refinement and improve clarity. All AI-assisted revisions were critically reviewed, edited, and approved by the authors, who remain fully responsible for the final content of the manuscript.

