## Supplementary material for "Smooth Curves, Similar Conclusions? Comparing Linear Regression and GAMLSS Neuropsychological Norms"

| **Supplementary Table 1.** Healthy control group demographics in previous normative studies | | | | | | |
| --- | --- | --- | --- | --- | --- | --- |
| **Study/test** | **n** | **Age** Mean (SD) [range] | **Years of education** Mean (SD) [range] | **Female** n (%) | **Raw score 1** Mean (SD) | **Raw score 2** Mean (SD) |
| CERAD WLT | 227 | 63.1 (8.6) [40-80] | 14.2 (3.2) [7-23] | 141 (62%) | Learning raw score 21.5 (3.3) | Delayed recall raw score 7.2 (2.0) |
| TMT | 292 | 62.9 (8.4) [41-84] | 13.2 (3.4)  [6-24] | 174 (59.6%) | TMT-A seconds  34.9 (11.1) | TMT-B seconds 82.4 (26.4) |
| FAS phonemic fluency | 204 | 62.8 (9.3) [40-84] | 14.0 (3.3) [8-23] | 121 (59.3%) | FAS total raw score 41.4 (11.0) |  |
| VOSP Silhouettes | 291 | 63.0 (68.4) [41-84] | 13.3 (3.3) [6-24] | 176 (60.5%) | VOSP Silhouettes raw score 22.3 (4.0) |  |
| *Notes.* n = number of participants; SD = standard deviation; CERAD WLT = Consortium to Establish a Registry for Alzheimer’s Disease Word List Test; TMT = Trail Making Test; FAS = letters F, A, and S phonemic fluency; VOSP = Visual Object and Space Perception Battery. | | | | | | |

| **Supplementary Table 2.** Final normative GAMLSS model coefficients | | | | | | |
| --- | --- | --- | --- | --- | --- | --- |
| Test | Parameter | Term | Estimate | SE | t | p |
| CERAD WLT  Learning | μ | (Intercept) | 1.768 | 0.328 | 5.39 | < .001 |
|  | μ | age | -0.021 | 0.004 | -4.99 | < .001 |
|  | μ | gender | 0.229 | 0.070 | 3.28 | 0.001 |
|  | μ | edu_years | 0.023 | 0.011 | 2.19 | 0.029 |
|  | σ | (Intercept) | -3.908 | 0.257 | -15.22 | < .001 |
| CERAD WLT  Delayed Recall | μ | (Intercept) | 2.363 | 0.611 | 3.87 | < .001 |
|  | μ | age | -0.035 | 0.008 | -4.59 | < .001 |
|  | μ | gender | 0.400 | 0.126 | 3.17 | 0.002 |
|  | μ | edu_years | 0.043 | 0.019 | 2.24 | 0.026 |
|  | σ | (Intercept) | -2.493 | 0.229 | -10.90 | < .001 |
| TMT-A | μ | (Intercept) | 2.544 | 0.131 | 19.39 | < .001 |
|  | μ | age | 0.015 | 0.002 | 7.25 | < .001 |
|  | σ | (Intercept) | -1.290 | 0.037 | -34.77 | < .001 |
|  | ν | (Intercept) | -0.335 | 0.179 | -1.86 | 0.063 |
|  | τ | (Intercept) | 1.015 | 0.157 | 6.46 | < .001 |
| TMT-B | μ | (Intercept) | 30.697 | 10.539 | 2.91 | 0.004 |
|  | μ | age | 1.077 | 0.136 | 7.91 | < .001 |
|  | μ | edu_years | -1.517 | 0.341 | -4.45 | < .001 |
|  | σ | (Intercept) | -1.300 | 0.037 | -34.91 | < .001 |
|  | ν | (Intercept) | -0.364 | 0.174 | -2.10 | 0.037 |
|  | τ | (Intercept) | 1.012 | 0.171 | 5.93 | < .001 |
| FAS  Phonemic Fluency | μ | (Intercept) | 21.810 | 3.042 | 7.17 | < .001 |
|  | μ | edu_years | 1.389 | 0.220 | 6.33 | < .001 |
|  | σ | (Intercept) | -1.388 | 0.053 | -26.16 | < .001 |
|  | ν | (Intercept) | 0.828 | 0.231 | 3.58 | < .001 |
| VOSP Silhouettes | μ | (Intercept) | 2.218 | 0.314 | 7.07 | < .001 |
|  | μ | age | -0.018 | 0.005 | -3.73 | < .001 |
|  | σ | (Intercept) | -2.764 | 0.132 | -20.92 | < .001 |
| *Notes.* μ = mu (location); σ = sigma (scale); ν = nu (skewness); τ = tau (kurtosis); SE = standard error; t = t-statistic; p = p-value; CERAD WLT = Consortium to Establish a Registry for Alzheimer’s Disease Word List Test; TMT = Trail Making Test; FAS = phonemic fluency (letters F, A, and S); VOSP = Visual Object and Space Perception Battery. Final models were fitted using beta-binomial distributions for both CERAD WLT subtests and VOSP Silhouettes, a Box-Cox Power Exponential Original (BCPEo) distribution for TMT-A, a Box-Cox Power Exponential (BCPE) distribution for TMT-B, and a Box-Cox Cole and Green (BCCG) distribution for FAS phonemic fluency. | | | | | | |

| **Supplementary Table 3.** GAIC comparisons between selected GAMLSS distributions and Gaussian models | | | | |
| --- | --- | --- | --- | --- |
| Test | Model | df | GAIC | ΔGAIC |
| CERAD WLT Delayed Recall | Beta-binomial (BB) | 5 | 892.06 | 0.00 |
| CERAD WLT Delayed Recall | Gaussian (NO) | 5 | 923.09 | 31.03 |
| CERAD WLT Learning | Beta-binomial (BB) | 5 | 1,158.46 | 0.00 |
| CERAD WLT Learning | Gaussian (NO) | 5 | 1,162.72 | 4.26 |
| FAS Phonemic Fluency | Box-Cox Cole and Green (BCCG) | 4 | 1,534.87 | 0.00 |
| FAS Phonemic Fluency | Gaussian (NO) | 3 | 1,535.80 | 0.93 |
| TMT-A | Box-Cox Power Exponential Original (BCPEo) | 5 | 2,129.45 | 0.00 |
| TMT-A | Gaussian (NO) | 3 | 2,191.65 | 62.20 |
| TMT-B | Box-Cox Power Exponential (BCPE) | 6 | 2,627.17 | 0.00 |
| TMT-B | Gaussian (NO) | 4 | 2,685.38 | 58.21 |
| VOSP Silhouettes | Beta-binomial (BB) | 3 | 1,610.48 | 0.00 |
| VOSP Silhouettes | Gaussian (NO) | 3 | 1,617.79 | 7.31 |
| *Notes.* GAIC = Generalized Akaike Information Criterion; df = degrees of freedom; ΔGAIC = difference from the best-fitting model within each test; CERAD WLT = Consortium to Establish a Registry for Alzheimer’s Disease Word List Test; TMT = Trail Making Test; FAS = phonemic fluency (letters F, A, and S); VOSP = Visual Object and Space Perception Battery; BB = beta-binomial; BCPEo = Box-Cox Power Exponential Original; BCPE = Box-Cox Power Exponential; BCCG = Box-Cox Cole and Green; NO = Gaussian distribution. | | | | |

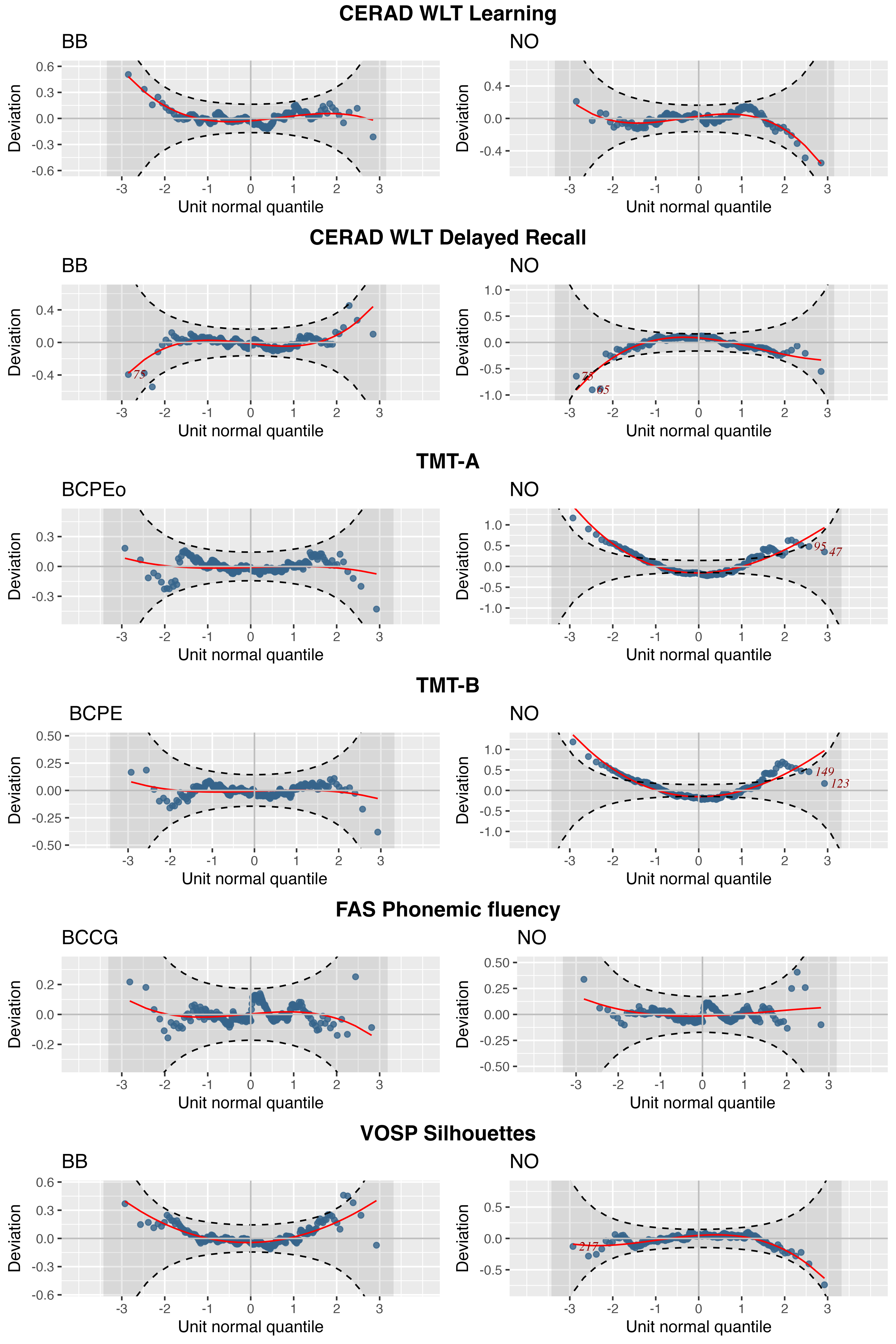

Supplementary figure 1. Worm plots for the selected Generalized Additive Models for Location, Scale and Shape (GAMLSS) distributions and corresponding Gaussian comparison models for each cognitive test. For each test, the selected distribution is shown alongside a Gaussian (NO) model fitted using the same predictor structure. Plot titles indicate the fitted distribution. Worm plots were used to visually assess model fit across the score distribution, with systematic deviations from the horizontal reference line indicating potential model misfit. BB = beta-binomial; BCPEo = Box-Cox Power Exponential Original; BCPE = Box-Cox Power Exponential; BCCG = Box-Cox Cole and Green; NO = Gaussian distribution. CERAD WLT = Consortium to Establish a Registry for Alzheimer's Disease Word List Test; TMT = Trail Making Test; FAS = phonemic fluency (letters F, A, and S); VOSP = Visual Object and Space Perception Battery.
